# Non-invasive glucose measurements in humans with time-gated mid-IR optoacoustic spectroscopy

**DOI:** 10.1101/2025.03.12.25323511

**Authors:** Alexander Prebeck, Uli Stahl, Maximilian Koch, Vasilis Ntziachristos

**Affiliations:** Chair of Biological Imaging, Central Institute for Translational Cancer Research (TranslaTUM), School of Medicine and Health & School of Computation, Information and Technology, Technical University of Munich, Munich, Germany; Institute of Biological and Medical Imaging, Bioengineering Center, Helmholtz Zentrum München, Neuherberg, Germany; Munich Institute of Biomedical Engineering (MIBE), Technical University of Munich, Garching b. München, Germany

**Keywords:** Diabetes management, photoacoustic spectroscopy, glucose sensing, health management, photoacoustic sensing, optoacoustic sensing

## Abstract

Frequent measurements of blood sugar are essential for the management of diabetes. While finger pricking offers accurate measurements of blood glucose, it is a procedure that causes discomfort and risk of infection. Conversely, minimally invasive biochemical sensors based on micro-needles do not assess glucose in blood but in the interstitial fluid. While most optical sensors also detect in bulk from the interstitial fluid, a depth-gated mid-infrared optoacoustic sensor (DIROS) was recently proposed to non-invasively detect glucose concentrations in blood by means of time-gating. While DIROS was previously demonstrated only in animals, herein we present the first pilot investigation of the sensor in humans, based on a multivariate model fit to measurement data obtained from healthy volunteers (n=5) during an oral glucose tolerance test. By time-gating optoacoustic signals, i.e. selecting time points corresponding to different depths within the skin based on the ultrasound time-of-flight, we confirm in humans an improved measurement accuracy when targeting deeper skin layers, which are rich in vasculature. The results set the first milestone towards depth-dependent in-blood glucose detection in humans and highlight potential for DIROS in clinical application.

## Introduction

Advancing blood glucose monitoring in humans, beyond the invasive extraction of blood by finger pricking, is of high interest due to the increasing prevalence of diabetes worldwide. Such measurements may also more broadly link to other applications, such as sports and wellness areas. Cutaneous, minimally invasive electrochemical micro-needles are considered today as a next step beyond the finger prick method, but measure glucose that diffused from blood capillaries into the interstitial volume, not directly in blood. However, this diffusion occurs in a delayed fashion and with a dilution factor that may vary with time, which may challenge the precision of the measurement. [1–7]

Moving beyond the use of minimally invasive electrochemical devices, non-invasive glucose sensing measures relative changes of different types of energy, such as electricity, light or THz radiation, interacting with glucose *in-vivo*, as the means for characterizing its concentration [8–14]. Such methods may leave the skin barrier intact but also generally offer measurements in bulk and sample glucose in volumes that mostly consist of the interstitial fluid (ISF). Moreover, methods based on energy interaction with tissue may be challenged by non-specific interactions of other biomolecules and skin structures. Of particular interest are methods using mid-infrared (MIR) radiation, which in contrast to approaches using light at other excitation wavelengths, e.g. Raman techniques or near-infrared (NIR) spectroscopy, offer both high detection sensitivity and high spectral specificity [8,13]. We have recently shown glucose sensing that uses optoacoustic (OptA) spectroscopy to enable depth-gated detection in the MIR. Termed, depth-gated mid-infrared optoacoustic sensing (DIROS), the approach was shown to improve the accuracy of non-invasive glucose measurements in the MIR, by selectively detecting signals at skin depths rich in vasculature [15]. Therefore, this technique goes beyond previous interrogations that used OptA detection to retrieve bulk measurements from tissues [16–23], i.e. to average all signals collected from the entire volume interrogated. In particular, DIROS was shown to improve the precision of the measurement when rejecting contributions from the superficial and metabolically inactive skin layers, thus selectively collecting signals only from the depths of the micro-vasculature at the epidermal-dermal junction. The findings corroborated imaging observations, which identified that superficial skin heterogeneity may yield variations of ∼30 % in the mean absolute deviation in cross-validation (MADCV) of an oral glucose tolerance test (OGTT), when performing measurements in bulk [23]. So far, no other method has used the time-of-flight feature of the OptA measurement to detect glucose as a function of depth [24–27].

Therefore, while DIROS was shown as a critical advance in non-invasive glucose monitoring, this original demonstration was performed only in mice. Mouse measurements are a fair indication of in-human performance, but it is nevertheless important to validate the technique in measurements on humans as well. Therefore, we aimed herein to provide a first investigation of DIROS application on the human skin and examine whether the improved performance of DIROS showcased in animals could be repeated in humans. To reach these goals, we obtained ethical approvals for the first *in-vivo* pilot demonstration of the technique in healthy volunteers (n=5). We employed time-gated OptA signals, during an OGTT, to select measurements that correspond to different depths within the skin by the acoustic time-of-flight. The time-gated measurements were processed, using a multivariate regression model fit, to investigate the relative performance when using different gates, against minimally invasive glucose measurements with a continuous glucose monitor (CGM), used as the gold standard. Our results confirm the previous observations of DIROS applied in animals and demonstrate improved accuracy in humans, when targeting skin layers rich in vasculature, over simple bulk measurements. We discuss how this pilot study is the first indication that DIROS could become the method of choice in non-invasive glucose measurements.

## Results

The DIROS sensor (**Fig.1a**) was implemented as a confocal arrangement of focused, pulsed MIR illumination (925/cm - 1250/cm) and a focused broadband piezoelectric ultrasound transducer (UST). The light pulses are focused through a zinc sulfide (ZnS) window to tissue placed between the window and the UST, and generate an ultrasound wave via the OptA effect. The ultrasound wave then propagates to the UST in trans-illumination mode. By wavelength scanning, DIROS detects a spectrum. Measurements of glucose-water solutions within the human physiological range (**Fig.1b**), exhibited the characteristic fingerprint peaks of glucose at 994/cm, 1080/cm, 1034/cm, 1108/cm and 1154/cm revealing the sensitivity of the system. Performing a leave-one-group-out (LOGO) cross-validation (CV) on spectra from a titration series like shown in Fig.1b, yielded a mean absolute error in cross-validation (MAECV) of 8.7 mg/dl (**Fig.1c**), which corroborates the high sensitivity of MIR-OptA spectroscopy to glucose concentrations, within the human physiological range, in the laboratory environment.

**Figure 1:**
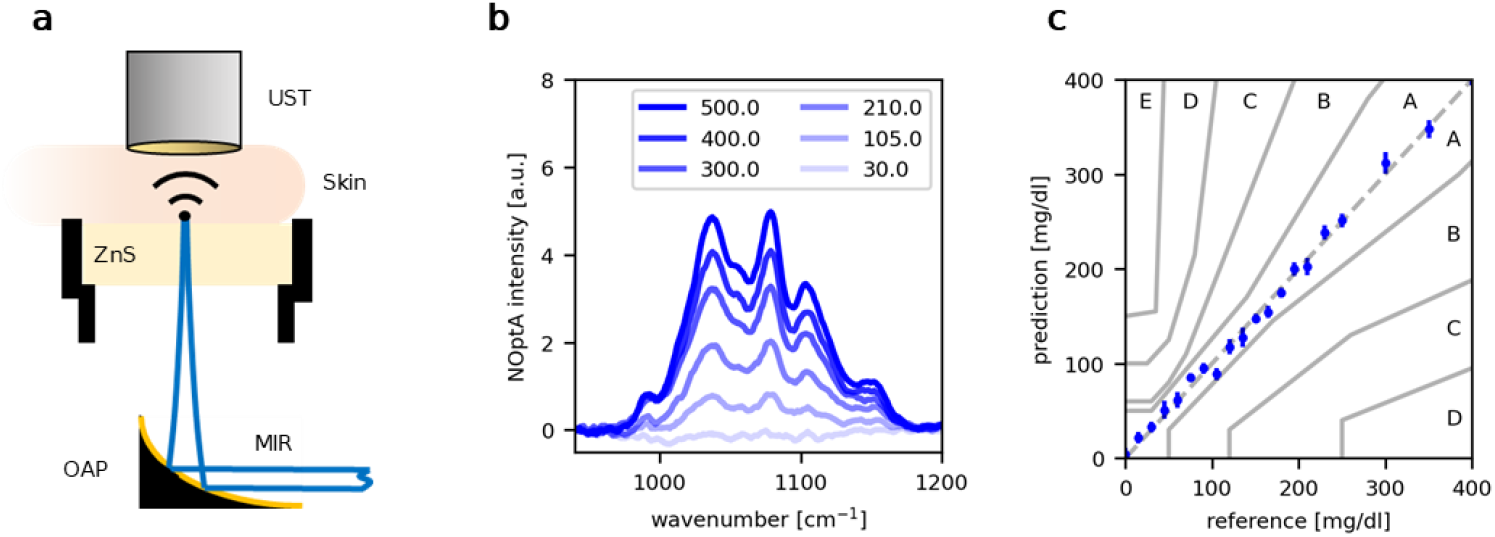
Schematic measurement principle of the depth-gated mid-infrared optoacoustic sensor (DIROS) used in this study as well as its sensitivity to glucose. (a) Schematic representation of the measurement principle used by DIROS, comprising of mid-infrared illumination (MIR) focused through a zinc sulfide window (ZnS) onto the skin by an off-axis parabolic mirror (OAP). A focused piezoelectric ultrasound transducer (UST) detects the pressure wave created by the optoacoustic (OptA) effect. (b) Normalized OptA (NOptA) spectra of glucose-water solutions with concentrations in the human physiological range recorded with the setup used in this study. (c) Results of a leave one group out cross-validation of a multivariate regression model when fit to spectra measured from glucose solutions within the human physiological range. Individual points show the mean and standard deviation of 15 measurements.

To evaluate the feasibility of non-invasive DIROS glucose measurements in humans, we performed a clinical pilot study with healthy volunteers (N=5). The study consisted of an OGTT as well as a control measurement, where only water was administered. DIROS measurements were acquired on the skin flap of the first interdigital space at the left hand of the volunteers (**Fig.2a-b**), in parallel to readings from a CGM that sampled glucose in ISF on the upper arm. Fig.2 shows the results of a leave one out cross-validation (LOOCV) performed on a multivariate regression model (see Methods) that was trained on the OptA spectra acquired during the OGTT.

**Figure 2:**
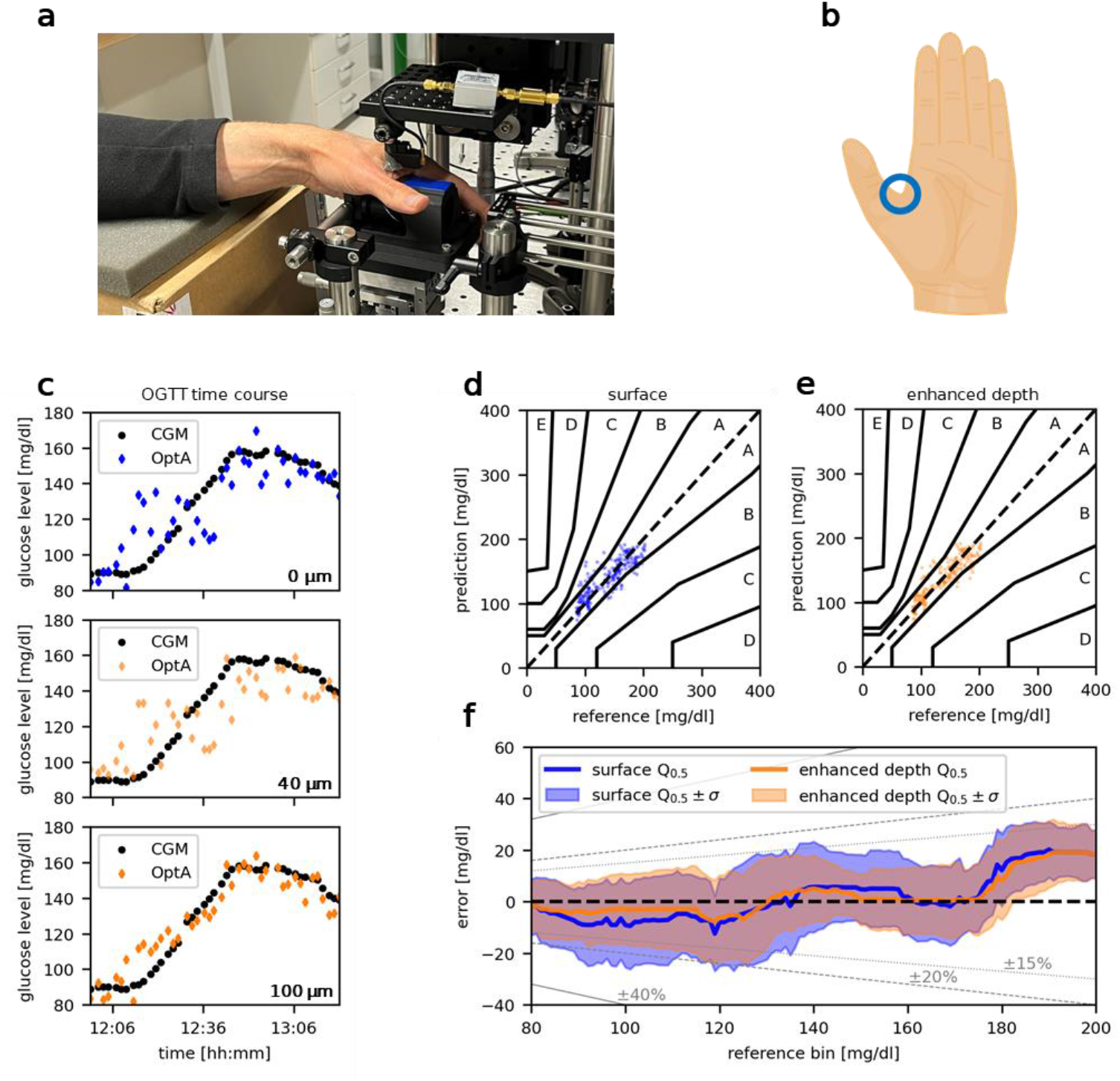
Summary of study results and overview of DIROS application in human measurements. (a) shows a photograph of a participant’s hand in the measurement setup. (b) marks the measurement location on the skin flap in the first interdigital space of the left hand. (c) shows exemplary time courses recorded by a continuous glucose monitor (CGM) as well as the measurement setup (OptA) during an OGTT when targeting different depths within the skin. (d-e) show the results from all participants in a single consensus error grid for measurements on the skin surface (d) as well as at an individually determined enhanced measurement depth (e). (f) shows the distribution of the measurement error as a function of reference value for measurements from the skin surface as well as from an individually determined enhanced measurement depth. The bold lines correspond to the median Q_0.5_, the contour lines to the standard deviation σ calculated within a sliding window of 20mg/dl width. Gray lines show error corridors of +-15%, +-20% and +-40% relative to the reference value.

We performed a comparative study to investigate the influence of time-gated detection on the accuracy of glucose measurement during an OGTT by performing a LOOCV analysis with and without time-gated measurements. Fig.2c shows exemplary LOOCV results for a single participant when we used spectra from different effective measurement depths from 0 µm up to 100 µm. For this participant, choosing a time-gate corresponding to deeper layers within the skin visibly improves the measurement results. Fig.2d-e on the other hand provide an overview of how time-gated detection influences the measurement results of all participants by performing a Consensus Error Grid (CEG) analysis [28]. Individual results for single participants range from 6.4 mg/dl to 16.7 mg/dl in MAECV. Measuring at the skin surface yields a cumulative MAECV of 13 mg/dl corresponding to a mean absolute percentage error (MAPE) of 10 %. Measuring at an individually enhanced depth reduces the overall MAECV to 11 mg/dl (MAPE: 8 %). At the skin surface the CEG analysis shows 89 % of measurement points in zone A and 11 % in zone B. At an enhanced depth, 91 % of points are found in zone A and 9 % in zone B. In both cases >99 % of the measurement points are found in zone A and B. The relative improvement with time-gated detection varies significantly between participants, with some measurements showing little to no improvement, while others show improvements of up to 30 % in MAECV.

Fig.2f shows, how the measurement error is distributed by calculating the median and standard deviation of measurement error within a 20 mg/dl wide sliding window. For both modalities, measuring at the skin surface or at an individually enhanced measurement depth, the measurement error is centered on 0 mg/dl up to reference values of roughly 180 mg/dl, where a stronger under-estimation of glucose measurements can be observed. Comparing time-gated measurements with surface measurements reveals that time-gated measurements show strongest improvement around reference values of 100 mg/dl, where measurement accuracy in the later application is needed the most. Additionally, time-gated detection increases the fraction of measurement points inside the 15 % and 20 % error corridors. The overall distribution of measurement error is significantly tighter (shows lower variance) when using time-gated detection as compared to surface detection. (Levene test for difference in variance, p=0.047)

## Discussion

We demonstrated the first DIROS pilot study in humans. We found that depth-gated OptA sensing in the mid-infrared could accurately measure glucose levels in humans, at precision levels similar to the ones achieved with the current state of the art. Importantly, we observed that as was the case for mouse measurements, time-gated detection from layers rich in microvasculature appears to improve the accuracy of DIROS and its agreement with minimally invasive CGM. Nevertheless, we do not expect a perfect agreement, since CGM only reported on ISF glucose values, which appear in a delayed fashion, compared to blood glucose fluctuations.

Unlike other sensors, DIROS is able to measure glucose at specified depths in human skin. By selecting depths abundant with blood vessels, the sensor enabled a more precise determination of glucose levels. This performance is a marked deviation to all optical techniques considered. Due to photon diffusion, pure optical methods only sample the entire volume that photons have diffused in, therefore sampling also from non-specific layers. Moreover, since light is attenuated as a function of depth, the stronger signals are produced from the metabolically mostly inactive stratum corneum [30], and other superficial skin layers that contain molecules that can contaminate the measurement. This bulk and surface-weighted operation of optical measurements may compromise the sensitivity and the accuracy of conventional optical sensors [8–10,12–14]. It has been also noted that non-glucose spectral contributions from the stratum corneum reduce the sensitivity and specificity of the measurements, especially in non-time-gated MIR recordings [16–23]. Machine learning methods have been developed to account for these unwanted contributions, but appropriate solutions to these problems remain challenging, especially in regard to ascertain if the computational approach chosen provides an appropriate solution, or if it merely produces lower error rates by overfitting specific datasets. [16,17,34]. DIROS avoids these issues and provides data that contains more accurate information, due to the fundamental improvement of reading only from selected layers and rejecting information from others. This signal selection, using the ultrasound time of flight, is already commonly used for 3D image formation in OptA imaging modalities, and can be translated to glucose measurements as we show in this study.

A key difference between the DIROS mouse measurements [15] and the study herein is that, unlike mice, the layer of the human skin that resulted in optimal performance had to be individually determined for each participant. This difference can be explained by the greater heterogeneity in skin structure expected in humans, over controlled mouse lines [31,35,36]. For the purposes of this pilot study, this layer determination was based on a brute force approach, calculating results for every depth and subsequently choosing the depth that yielded the lowest MAECV for each participant. However, in the future, this depth selection process could be automated by localizing prominent blood chromophores like hemoglobin in parallel with DIROS, making it suitable to identify the depth of the vascular layer in the epidermal-dermal junction of each individual.

The pilot study indicated that compared to industry standards DIROS already seems very promising. Considering the boundaries set for the distribution of measurement errors by ISO15197 for in vitro glucose measurement devices, DIROS results in this study already show a partial match with 100 % of measurement results in the CEG zones A and B. Furthermore, to gage DIROS’ performance in the context of a potential continuous glucose monitoring device, we compared our measurement results against the accuracy requirements stated by the American Food and Drug Administration (FDA) for integrated continuous glucose monitoring (iCGM) devices (FDA 21CFR862.1355 [29]). Considering the FDA’s conditions A-I and their respective allowed distributions of measurement error, e.g. the fraction of measurement points within the error corridors shown in Fig.2f, DIROS error distributions lie within the borders specified wherever measurements were recorded in this study. However, these results provide only rough comparisons, as we used CGM data as reference measurements in this study, rather than capillary blood tests.

The selection of a CGM as the validation method herein was guided by the need to provide multiple measurements over the course of the OGTT, which is challenging with the finger pricking method. Consequently, we followed the recommendation of the Ethics Committee to use CGM over finger pricking for this pilot study. Nevertheless, the CGM study is known to contain errors, with a MARD of 7.8 % reported in the literature [37]. Taking into account DIROS’ MAPE of 8 % and the CGM’s MARD of 7.8 %, DIROS is likely to conform with the FDA’s requirement for iCGMs, stating in one condition only ±20 % accuracy for at least 87% of measurements across the entire measurement range [29].

Moreover, the measurements herein were conducted on the easily accessible interdigital skin-flap, as shown in Fig. 2b. Alternative sites, such as the earlobe or volar forearm might lead to higher observation accuracy, as they are reported to be less prone to sweating and have a more favorable skin structure with thinner stratum corneum layers [21–23]. This suggests that DIROS performance could be further enhanced by optimizing the DIROS location. In the long term, transitioning from a trans-illumination to an epi-illumination OptA setup would allow measurements on any skin surface. The next step in the DIROS development is to recruit a larger number of volunteers and employ the data to research a global regression model, aiming to avoid the need for calibration. A study in a larger cohort would also enable interrogation of the effects of population variability and confounding factors such as age or gender on the measurement.

DIROS could also be adapted to measuring other relevant metabolites that exhibit distinct absorption features in the MIR wavelength region, such as lactate. With the potential for miniaturization and scalable production, DIROS could be integrated into wearable devices for real-time measurements that will allow rapid interventions, for example in response to changes in blood glucose levels, as is possible now with CGMs. Such wearable devices would also be desirable to the broad consumer market that is increasingly concerned with performance in sport and in health monitoring.

## Methods

### Experimental Setup

The light source used for the experiments in this study is an external cavity quantum cascade laser (EC-QCL) from Daylight Solutions with a tuning range of 925/cm to 1250/cm. The beam is first expanded by a 6x reflective beam expander and then focused by an off-axis parabolic mirror (OAP) with a focal length of 1 inch. This results in a theoretical diffraction limited spot size of roughly 30 µm in diameter. The optical focus is placed on one surface of a ZnS window with a MIR antireflective coating. A broadband focused piezoelectric UST (f_center_ = 25 MHz, piezocomposite) is confocally aligned to the MIR excitation. The signal from the UST is amplified in two stages by 30 dB and 40 dB respectively. After amplification, the signal is digitized using a PCI oscilloscope card (GaGe). For skin safe measurements, the setup’s output power is adjusted to stay within maximum permissible exposure values, specified by relevant German legislation (TROS Laserstrahlung). The wavelength of the QCL is tuned in 10 nm steps from 8 µm to 10.84 µm to acquire spectra, resulting in 285 wavenumbers per spectrum. For every wavenumber, 5000 raw OptA transients are acquired and averaged to reduce uncorrelated in-band noise. A Butterworth filter (order 6, f_-3dB_ = 30 MHz) is applied digitally to reduce out-of-band noise.

### In-vivo measurements

The in-vivo experiments were performed in a pilot clinical study approved by the TUM Ethics Commission (registered with the German Clinical Trials Register: DRKS00036371). Participants (male: 4, female: 1. age: mean 42 y, standard deviation 11 y) arrived for measurement after a 12-hour fast. Participants sat relaxed on a chair, put their lower arm on a foam pad and placed their hand on the measurement interface. The transducer’s cavity was filled with distilled water, enclosed by a latex membrane and coupled to the participant’s hand using a thin layer of ultrasound gel. The transducer’s alignment to the optical excitation was subsequently adjusted to achieve optimal signal amplitude and the sample was shifted using linear stages to select a measurement point with a defined acoustic interface to the ZnS window.

DIROS and CGM (Freestyle Libre 3, Abbott) readings were taken continuously during the measurement. During the first 10-15 minutes, a background measurement was conducted. Subsequently, depending on whether it was an OGTT or a control measurement, participants were given 300 ml of water either with or without 75 g of glucose mixed in. Participants were instructed to consume the solution within a maximum of 5 minutes. From the time of glucose administration measurements continued for approximately 90 minutes.

### Time-gated optoacoustic detection

To selectively measure deeper inside the skin, OptA spectra were generated by applying a specific time gate to the Hilbert envelope of the OptA transients, effectively selecting a measurement depth using the acoustic time-of-flight. The specific method used by DIROS can be found elsewhere [15].

### Data Analysis

The spectra generated by DIROS during the in-vivo measurements were analyzed by use of a multivariate regression model. Before analysis, wavenumbers of low laser output power were removed. Furthermore, a local outlier factor method (Python sklearn) discarded spectra showing motion artefacts. After feature standardization and decorrelation using principal component analysis, a Bayesian ridge regressor (Python sklearn) was fit to the data. This approach is similar to principal component regression, a method well-suited for chemical and spectral analysis.

For every measurement series, the model was individually fit to the spectra of all time-gates and evaluated by LOOCV using the mean absolute error in cross-validation (MAECV) as a metric. Subsequently, the time-gate that resulted in the lowest MAECV for a measurement series was individually selected. The depths evaluated, calculated from the speed of sound in soft tissue (1500 m/s), reach from 0 µm in 20 µm steps up to 100 µm in depth.

## Data Availability

All data produced in the present study are available upon reasonable request to the authors

## Abbreviations

AR: Acoustic Resonance
CEG: Consensus Error Grid
CGM: Continuous Glucose Monitor
CV: Cross-Validation
DIROS: Depth-Gated Mid-Infrared Optoacoustic Sensor
EC-QCL: External Cavity Quantum Cascade Laser
FDA: American Food and Drug Association
iCGM: Integrated Continuous Glucose Monitor
ISF: Interstitial Fluid
LOGOCV: Leave-One-Group-Out-Cross-Validation
LOOCV: Leave-One-Out-Cross-Validation
MADCV: Mean Absolute Deviation in Cross-Validation
MAECV: Mean Absolute Error in Cross-Validation
MAPECV: Mean Absolute Percentage Error in Cross-Validation
MIR: Mid-Infrared
NIR: Near-Infrared
NOptA: Normalized Optoacoustic
OAP: Off Axis Parabolic Mirror
OGTT: Oral Glucose Tolerance Test
OptA: Optoacoustic
OptAS: Optoacoustic Spectroscopy
QCL: Quantum Cascade Laser
RMSECV: Root Mean Square Error in Cross-Validation
UST: Ultrasound Transducer
ZnS: Zinc Sulfide

## Acknowledgements

The research leading to these results was funded under the European Union’s Horizon 2020 and Horizon Europe research and innovation programme under grant agreement no. 862811 (RSENSE) and no. 101058111 (GLUMON). We thank Dr. Serene Lee for her attentive reading and improvements of the manuscript.

## Competing Interests

V.N. is a founder and equity owner of Maurus OY, sThesis GmbH, iThera Medical GmbH, Spear UG and I3 Inc. M.K. is an equity owner and contractor for sThesis GmbH.

## Notes

### Clinical Trial

DRKS00036371

### Funding Statement

The research leading to these results was funded under the European Unions Horizon 2020 and Horizon Europe research and innovation programme under grant agreement no. 862811 (RSENSE) and no. 101058111 (GLUMON).

### Author Declarations

The Ethics Commission of Technical University Munich gave ethical approval for this work (2023-390-S-NP).

